# Observational Report on Functional SARS-CoV-2 Immune Responses in Immune Competent and Compromised Individuals

**DOI:** 10.1101/2024.11.08.24316708

**Authors:** Laura J Peek, Alicia Patrick, Niki Givens, Brittany D’Alessio, Amanda Weaver, Leisa Jackson, Thomas J White, Gary A Pestano

## Abstract

**Objectives:** Durable immune responses against SARS-CoV-2 are generally observed in previously infected individuals. Immunocompromised individuals may show less durable responses. Investigation of immune responses is needed to inform individuals on whether measures such as boosters may be advisable.

**Methods:** Three SARS-CoV-2 ELISA tests were utilized to monitor responses in 82 individuals in the United States [nucleocapsid protein (N)-specific antibody and spike RBD (receptor binding domain)-specific neutralizing antibody (nAb), and spike-specific interferon-γ (IFN-γ) release by T cells]. De-identified case reports highlight results for this cohort; 21 previously infected and vaccinated, 57 never infected and vaccinated, 2 breakthrough infections post-vaccination and 2 neither infected nor vaccinated.

**Results:** The vaccines generated nAb responses in naïve donors, and in previously infected individuals with waning responses, immunosuppression, or weak infection responses. The cutoff for the nAb potency was established at 41.1 IU/mL when calibrated to the WHO International Standard (IU) using the NCI SARS-CoV-2 Standard. This relationship has the critical potential to allow comparisons of nAb measures to be utilized in studies of correlates of protection. Further, previously infected donors demonstrated robust T cell activity, while vaccinated and infection-naïve individuals had lower responses.

**Conclusions:** Calibrated ELISA tests may aid broad vaccination strategies given their low cost and wide accessibility. Additional studies that relate measures of the adaptive responses to immunity from serious disease and infection are needed.

## 1. Introduction

Immune responses against severe acute respiratory syndrome coronavirus 2 (SARS-CoV-2) have been observed in both previously infected and vaccinated individuals^1,2^. Reports suggest a vaccine efficacy of 91% (95% CI [88.8-93.0]) up to 6 months post vaccination for the BNT162b2 mRNA vaccine^3^, although specific and quantifiable correlates of protection and duration of such responses remain unknown^1,2^. However, many immunocompromised individuals, including patients with autoimmune disease, cancer patients, transplant recipients and the elderly, may mount a reduced immune response after vaccination and/or infection, leaving them uncertain of their protection^4,5^. Further investigation of the magnitude and duration of SARS-CoV-2 adaptive immune responses and their relationship to protective immunity is therefore needed to help inform the robustness of both B and T cell responses, timing of when the immune responses may be waning, and whether additional measures such as booster vaccinations may be advisable.

Here, we present representative de-identified case reports from an observational study of longitudinal surveillance of immune responses in individuals representing the general population in the United States. Twenty-one of the donors were previously infected with SARS-CoV-2 and subsequently vaccinated, 57 donors were never infected and received SARS-CoV-2 vaccination, 2 donors experienced breakthrough infections of SARS-CoV-2 after full vaccination, and 2 donors were neither infected nor vaccinated at the time of this report.

## 2. Materials and Methods

### 2.1 Study Cohort

The observational study (DROPLET, BDSX-CD-004) was approved by the Advarra Institutional Review Board (IRB) and all donors provided written informed consent. A statistical analysis plan (SAP) was not required for an observational study of all comers regardless of pre-existing immune conditions. We note that in an observational study of this nature certain biases will naturally exist, for instance in gender and age diversity, ethnicity, education, and occupations.

The eighty-two donors enrolled represent 21 infected and subsequently vaccinated, 57 vaccinated and never infected, 2 breakthrough infections after full vaccination and 2 controls neither vaccinated nor infected. Vaccinated donors represent 3 vaccines (mRNA-1273 [Moderna Inc.], BNT162b2 [Pfizer/BioNTech Inc.], Ad26.COV2.S [Johnson and Johnson]) authorized for emergency use by the US FDA at the initiation of this study. We note that BNT162b2 was approved for individuals ages 16 and up, as of 23 Aug 2021, and is marketed as Comirnaty.

All FDA EUA tests used in this study were evaluated in global harmonization studies aimed at elucidating correlations with the WHO and other international standards as are relevant^6^. Relational results are included herein for the cPass neutralization antibody to the Human SARS-CoV-2 Serology Standard from the Frederick National Laboratory for Cancer Research, Vaccine Immunity and Cancer Program (Lot Number COVID-NS01097), which is calibrated to the WHO International Standard.

### 2.2 Blood Collection & Testing Methods for Monitoring Adaptive Immune Responses

All adaptive immune monitoring tests were conducted in a standard BSL2 clinical laboratory using commonly available ELISA technology. Blood was drawn approximately once per month between May 2020 and August 2021 from each of the 82 enrolled donors, and anti-SARS-CoV-2 antibodies were measured by ELISA in a centralized CLIA laboratory (Biodesix, Inc.). B cell responses to different antigens in SARS-CoV-2 were evaluated using two commercially available FDA EUA ELISA test kits. Specifically, total nucleocapsid protein (N)-specific antibody (Platelia SARS-CoV-2 Total Ab Assay, catalog #12015289, Bio-Rad Inc.) and spike RBD (receptor binding domain)-specific neutralizing antibody (nAb) responses (cPass^TM^ Neutralization Antibody Detection Kit, catalog #L00847, GenScript Inc.) were measured at a single dilution using the manufacturers’ instructions for use.

Dilution series of the Human SARS-CoV-2 Serology Standard (Lot Number COVID-NS01097; NCI-Frederick National Laboratory for Cancer Research; Vaccine Immunity and Cancer Program), which is calibrated to the WHO International Standard, was also evaluated by both assays to allow future inter-assay and inter-laboratory comparisons. Given this study’s focus on clinical relevance of these assays, we focused our analyses on test results at the lower range since it is most meaningful to assess results at or near the threshold of positive and negative cutoff in the evaluation of biomarkers with potential clinical utility.

Additionally, 11 donors previously vaccinated, previously infected and subsequently vaccinated, or never vaccinated nor infected, were selected to assess specific interferon-γ (IFN-γ) release by T cells using a novel research use only (RUO) assay performed according to the manufacturer’s instructions for use (QuantiFERON^®^ SARS-CoV-2 Starter Set (catalog #626115), Control Set (catalog #626015) and IFN-γ ELISA (catalog #626410); Qiagen Inc)^7^.

#### 2.2.1 α-Nucleocapsid Total Antibody Test

The α-nucleocapsid (anti-N) total antibody test detects antibodies (IgG, IgM & IgA combined) to the highly abundant nucleocapsid protein. The assay is performed as a one-step antigen capture ELISA as described in the FDA EUA instructions for use for the Platelia SARS-CoV-2 Total Antibody test kit^8^. The diluted plasma (1:5), or Human SARS-CoV-2 Serology Standard (8 point, 1.5-fold dilution series, starting at 1:90 dilution), was mixed with SARS-CoV-2 nucleocapsid protein coupled with horseradish peroxidase (HRP) enzyme at a 1:1 ratio, and 100 μL added to a 96-well plate coated with the nucleocapsid protein. The plate was covered with an adhesive plate sealer and incubated at 37°C for 1 hour. The plate was then washed five times with the prepared Working Washing Solution provided in the kit, and 200 μL of the Enzyme Development Solution was added to each well. After a 30-minute incubation in the dark at room temperature (18-30°C), the reaction was stopped by adding 100 μL per well of an acidic stopping solution and mixing thoroughly before measuring the optical density (OD) at 450 nm using a spectrophotometer. The assay readout was a ratio of the specimen OD to cutoff control OD. A specimen-to-cutoff ratio ≥1.0 was declared positive, <0.8 was negative, and in between was reported equivocal with the recommendation of another specimen collected a few days later. Results were calculated as a percentage of the maximum measurable specimen-to-cutoff ratio of 6.0. Results measured above the reportable range of the assay were not further diluted, but rather reported as 100% for purposes of this study.

#### 2.2.2 α-RBD Neutralization Antibody Test

The α-RBD nAb test specifically measures a subset of antibodies that can block the interaction between the RBD on the SARS-CoV-2 spike protein and the human host receptor angiotensin-converting enzyme 2 (ACE2). The assay is performed as a blocking ELISA as described in the FDA EUA instructions for use in the cPass™ SARS-CoV-2 Neutralization Antibody Detection Kit^9^. The surrogate virus neutralization test (SVNT) cPass assay was clinically validated and shown to be 100% sensitive and specific when compared to a gold standard PRNT (plaque reduction neutralization test), with qualitative analysis results 100% in agreement.

In our laboratory, the diluted plasma (1:10), or Human SARS-CoV-2 Serology Standard (8 point, 2-fold dilution series, starting at 1:5 dilution), was pre-incubated 1:1 with RBD protein conjugated to HRP at 37°C for 30 minutes. The mixture (100 μL) was then added to a 96-well plate coated with human ACE2 receptor protein; the plate was sealed and incubated an additional 15 minutes at 37^°^C. The plate was washed four times with 260 μL/well Wash Solution provided in the kit before addition of 100 μL/well TMB (3,3’,5,5’-Tetramethylbenzidine) substrate that was allowed to react for 15 minutes at room temperature. Fifty microliters of an acidic stop solution were added to each well, and the OD was measured at 450 nm using a spectrophotometer. The nAb assay readout was percent signal inhibition by neutralizing antibodies, which was calculated to be the OD value of the sample relative to the OD of the negative control subtracted from one^7,9-11^. Results ≥30% signal inhibition were declared positive, and results <30% were reported negative based on previously conducted clinical validation studies^7^. The cPass assay has a current EUA for a qualitative and semi-quantitative interpretation of results^9^, and here we utilized the numeric percent signal inhibition (semi-quantitative measure) for comparing relative changes over time. We note that the current FDA EUA extended the cPASS test to result semi-quantitative results using a manufacturer developed calibration control^8^. However, these results are not comparable to other SARS-CoV-2 neutralizing (or binding) antibody assays since the performance characteristics of each SARS-CoV-2 antibody test is uniquely established^9^. To permit our results to be comparable to a gold standard reference, a dilution series of the Human SARS-CoV-2 Serology Standard, which is calibrated to the WHO International Standard, was used to calibrate the cPASS neutralizing antibody assay. Results measured above the reportable range of the assay were not further diluted, but instead are reported as 100% for purposes of this study.

#### 2.2.3 Interferon-γ Test for SPIKE RBD Specific T cell responses

An interferon-γ test was used to assess the specific T cell response to SARS-CoV-2 infection and/or vaccination in selected donors. In this exploratory study, SARS-CoV-2 blood collection tubes coated with specific SARS-CoV-2 peptides from spike antigens (S1, S2 RBD) were used to stimulate lymphocytes in 1 mL heparinized whole blood overnight (16-24 hours) at 37°C. After treatment of the blood samples, the IFN-γ levels were measured using the QuantiFERON IFN-γ ELISA kit, utilizing an 8-point IFN-γ standard curve. Plasma (50 μL) from the stimulated samples was added alongside the standard dilutions (prepared from the recombinant human IFN-γ standard provided in the kit) to the QuantiFERON 96-well plate containing 50 μL prepared conjugate, and the mixture was incubated for 2 hours at room temperature. After washing the plate 6X with 400 μL per well of the prepared Wash Buffer provided in the kit, 100 μL Enzyme Substrate Solution was added to each well and the sealed plate incubated for 30 minutes at room temperature. The reaction was stopped by addition of 50 μL Enzyme Stopping Solution before measuring the OD at 450 nm with background correction at 620 nm. The IFN-γ concentration (reported in International Units per ml [IU/ml]) was calculated from the line of best fit for the 8-point standard curve by regression analysis. QuantiFERON Mitogen and Nil tubes were also processed for each donor to serve as positive and negative assay and donor controls.

### 2.3 PCR testing for Diagnosis and Monitoring of SARS-CoV-2 Viral Infection

All donors were routinely tested for SARS-CoV-2 infection using a highly sensitive partition-based PCR method. The FDA EUA SARS-CoV-2 ddPCR (Droplet Digital PCR) Kit^12^ (catalog #12013743, Bio-Rad Inc., Hercules, CA) is an endpoint RT-PCR test intended for the detection of nucleic acids from SARS-CoV-2 in nasopharyngeal, oropharyngeal, anterior nasal and mid-turbinate nasal swabs, nasopharyngeal washes/aspirates and nasal aspirates, as well as bronchoalveolar lavage (BAL) specimens. The oligonucleotide primers and probes for detection of SARS-CoV-2 are the same as those reported by CDC and were selected from regions of the virus nucleocapsid (N) gene. The panel is designed for specific detection of the 2019-nCoV (two primer/probe sets). An additional primer/probe set to detect the human RNase P gene (RP), used as an internal control, is also included in the panel. The 2019-nCoV CDC ddPCR Triplex Probe Assay (catalog # 12008202, Bio-Rad Inc.) includes these three sets of primers/probes into a single assay multiplex to enable a one-well reaction.

Briefly, respiratory specimens were collected into molecular transport media (PrimeStore^®^ MTM; Cat #LH-PSMTM-5-50, Longhorn Vaccines and Diagnostics LLC, MD) and shipped to the Biodesix testing laboratory. RNA was extracted using Quick-RNA Viral 96 kit (Zymo Research, catalog #R1040). A human specimen control sample prepared from A549 cells was processed with each batch as an extraction control. Three hundred microliters of the transport media was mixed with 300 µL inactivation solution (DNA/RNA Shield catalog number R1200-125, Zymo Research). Two hundred microliters of the sample/shield mixture was combined with 400 µL Viral RNA buffer and applied to a 96-well spin column plate. The plate was centrifuged for 5 minutes at 2,200 x g in a Heraeus Megafuge 8 centrifuge (Thermo Scientific). The columns were washed twice with 500 µL Viral wash buffer and once with 100% ethanol; after application of each wash, the plate was centrifuged for 5 minutes at 2,200 x g and the flow-through discarded. The plate was then spun for an additional 2 minutes at 2200 x g to dry the columns. Forty microliters nuclease-free water was applied to each column, and the plate was centrifuged for 5 minutes at 2,200 x g to elute the RNA. The RNA was held on ice until use in ddPCR followed by storage in ultra-low freezer. We additionally performed single column RNA extractions (Zymo Research, D4014) on some specimens when full batches of specimens were not available for testing. In these cases, all volumes were the same, but centrifugation speeds and times differed slightly. Single columns were spun at 10,000 x g for all steps: 2 minutes for binding, 30 seconds for each of the washes, and 2 minutes for the drying spin. Purified RNA was eluted into 1.5 mL tubes and held on ice until use in ddPCR followed by storage in ultra-low freezer.

The PCR reaction mix comprised 5.5 µL RNA and 16 µL PCR master mix (Bio-Rad One-Step RT-ddPCR Advanced Kit for Probes, catalog number 1864021). Twenty microliters of the PCR reaction mix were used to generate droplets on a QX200 Droplet Generator (Bio-Rad Inc.). A ddPCR positive (SARS-CoV-2 Standard, Exact Diagnostics, catalog number COV019) and negative control (no template) were processed with each batch. The droplets were transferred to a 96-well PCR plate and run on a combined RT-ddPCR thermocycling program (Bio-Rad model S1000 or C1000 Touch). After thermocycling, the plate was transferred to a QX200 droplet reader (Bio-Rad Inc.). The results from the reader were analyzed to determine copy numbers of N1, N2, and RPP30 detected in each 20 µL PCR. Results were generated based on thresholds for each target using QuantaSoft version 1.7.4.0917 (Bio-Rad Inc.). Variant testing was performed with the same method using variant specific assays (Bio-Rad Inc.; K417N dMDS817055273 and L452R dMDS983315944).

## 3. Results

Of the 82 donors consented for the study, 40 (49%) underwent surveillance testing for more than 4 months, with 17 donors having more than 6 months surveillance and 4 donors with more than 1 year (maximum of 14 months).

Of the 21 donors vaccinated and previously infected, 17/21 (81%) had more than 4-months surveillance. Twenty-two of the 57 (39%) vaccinated and never infected had more than 4-months surveillance. Both breakthrough cases were confirmed to be DELTA variant positive (Bio-Rad Inc.; K417N dMDS817055273 and L452R dMDS983315944) by conducting variant-specific ddPCR on nasal swab specimens. These two cases were enrolled into the study late and therefore only had a single measurement. The two negative control donors (neither vaccinated nor infected) undergo weekly ddPCR testing and have participated in the surveillance study for at least 3 months and as long as 5 months.

### 3.1 Antibody Responses

In 57 donors without previous SARS-CoV-2 infection, positive nAb responses were generally observed (n = 53/57; i.e., 93% with signal inhibition ≥30%^10,11^) post full vaccination, and negative for anti-N antibodies. As expected, the levels and duration of response varied (representative cases in **Figure 1**). Of the four donors who did not have a positive nAb response to vaccination, three of the four had known conditions associated with immune suppression (e.g., rheumatoid arthritis, multiple myeloma, etc), and the fourth donor (donor V-1, **Figure 1**) remained negative for nAbs nearly 2.5 months after receiving the J&J vaccine (Ad26.COV2.S). The latter donor subsequently received a dose of the Pfizer vaccine (BNT162b2/Comirnaty), and 14 days later tested positive for nAbs. All 6 donors not previously infected who received the J&J vaccine showed lower nAb levels as compared to those vaccinated with the Moderna or Pfizer vaccines. Regarding duration of nAb response, there is evidence of waning nAb levels in a few donors (**Figure 1**), the rate of which is variable; in fact, donor V-5 (**Figure 1**) crossed the nAb positivity threshold to become negative just ∼4.5 months post-vaccination, but nAb levels continued to be detectable.

**Figure 1.**
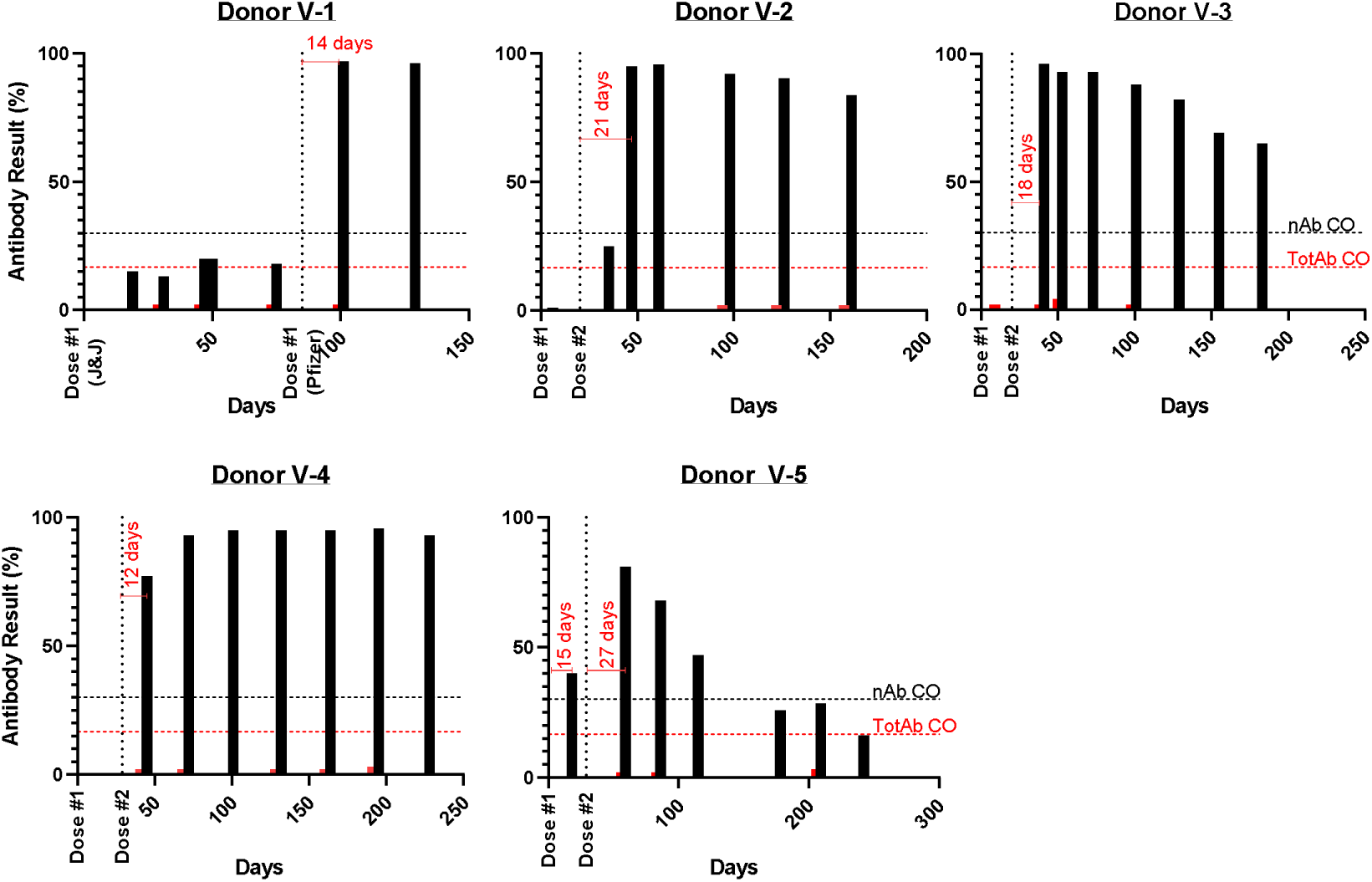
Measurements of SARS-CoV-2 total antibodies and neutralizing antibodies in select case study donors who were vaccinated but not previously infected with SARS-CoV-2. Neutralizing antibody levels (black bars) are reported as % signal inhibition (quenching assay) with levels ≥30% indicating a positive result [9,10] as shown by the black horizontal dashed line (nAb CO), whereas the specimen-to-cutoff ratio for the total antibody test (red bars) has been converted to % of maximum measurable specimen-to-cutoff ratio of 6.0. The cutoff for the total Ab test is shown by the red horizontal dashed line (TotAb positive CO). Abbreviations: Ab, antibody; CO, positive cutoff; nAb, neutralizing antibody; TotAb, total antibody.

In 21 donors previously infected with SARS-CoV-2, the immune response was variable with most donors maintaining nAb levels ≥30% and positivity for anti-N antibody levels prior to vaccination (**Figure 2**). Two donors in this category (2/21 = 10%), including donor I-6, demonstrated consistently negative nAb levels prior to vaccination (**Figure 2**). Previously infected donors exhibited increases in nAb levels following immunization (n = 21/21; i.e., representative cases in **Figure 2**). Of note, Donor I-1 was taking an immunosuppressant and experienced waning of both anti-N antibodies and nAb. Following vaccination of this donor, nAb levels were significantly boosted whereas anti-N antibodies remained minimal (**Figure 2**). Neutralizing antibody levels appear to persist for a longer duration as compared to anti-N antibody levels, which tend to wane in some donors (e.g., I-1, I-4, I-6, I-7, I-11, I-12, I-13 & I-14); however, other donors (e.g., I-3, I-9 & I-10) have anti-N levels that have remained fairly consistent across more than 6-12+ months. It should also be noted that the anti-N levels vary quite dramatically across previously-infected donors, with some donors only producing very low levels (e.g., donor I-6, I-12), others with moderate levels (e.g., donor I-5, I-7, I-11) and several donors with quite high levels (e.g., donors I-2, I-3, I-9) above the reportable range of the assay at the 1:5 specimen dilution tested.

**Figure 2.**
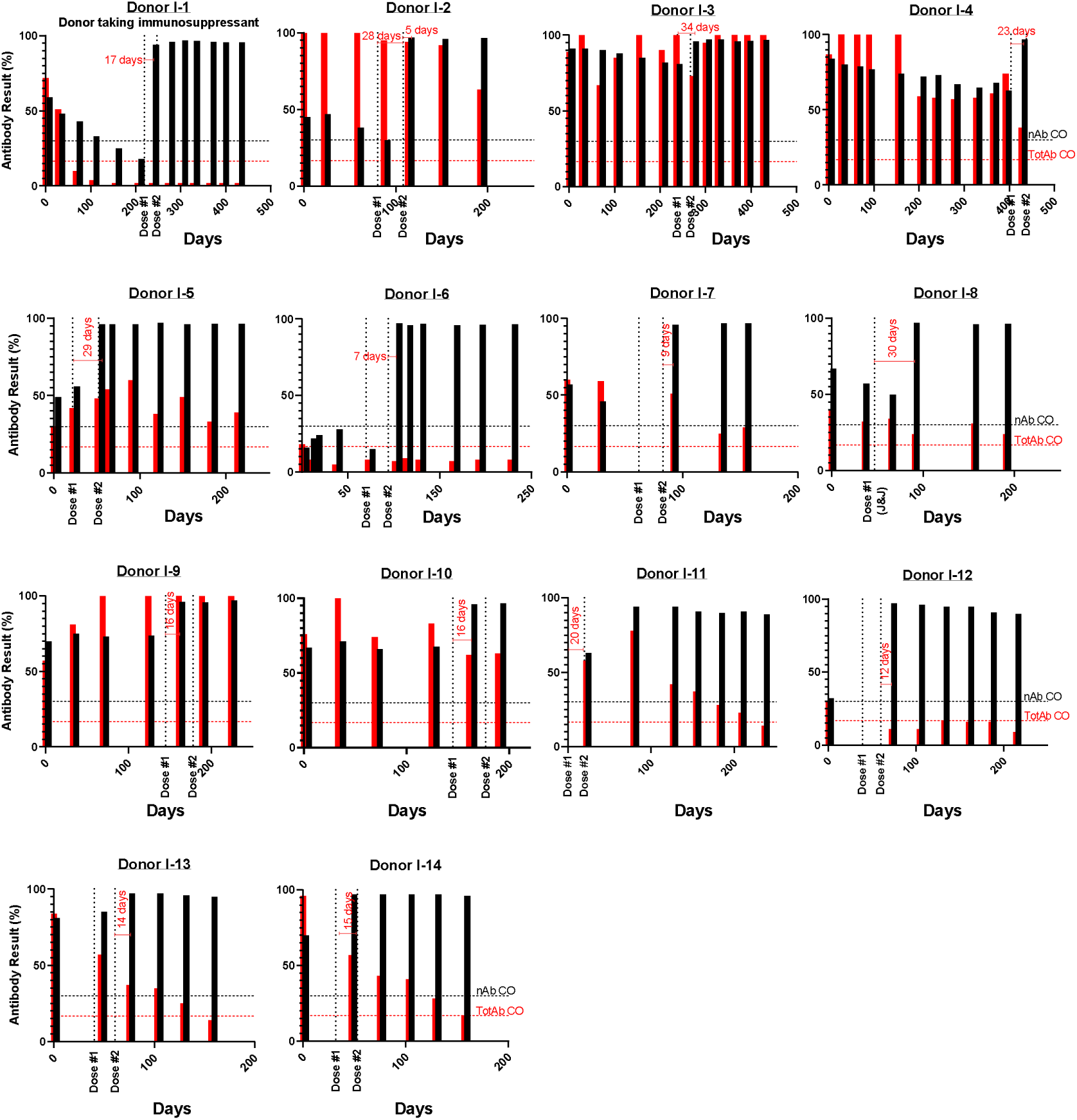
Measurements total antibodies and neutralizing antibodies in select case study donors who were infected with SARS-CoV-2 and subsequently vaccinated. Neutralizing antibody levels (black bars) are reported as % signal inhibition (quenching assay) with levels ≥30% indicating a positive result [9,10] as shown by the black horizontal dashed line (nAb CO), whereas the specimen-to-cutoff ratio for the total antibody test (red bars) has been converted to % of maximum measurable specimen-to-cutoff ratio of 6.0. The cutoff for the total Ab test is shown by the red horizontal dashed line (TotAb CO). Abbreviations: Ab, antibody; CO, positive cutoff; nAb, neutralizing antibody; TotAb, total antibody.

The 2 donors who experienced breakthrough cases after vaccination were enrolled post-infection. One donor tested positive for SARS-CoV-2 infection 39 days after receiving the second dose of vaccine and experienced no symptoms; nAb levels were high (89% signal inhibition), likely due to both vaccination and infection, while anti-N antibody levels remained very low (2% of the maximum) when tested 2 months after infection. The other donor had a persistent cough and tested positive for SARS-CoV-2 infection 6 months after vaccination; both nAb and anti-N antibody levels were high (95% signal inhibition for nAb and the anti-N result was above the reportable range of the assay) when tested 2.5 weeks after infection.

The two donors who were neither infected nor vaccinated to date had negative antibody levels throughout (**Figure 3**).

**Figure 3.**
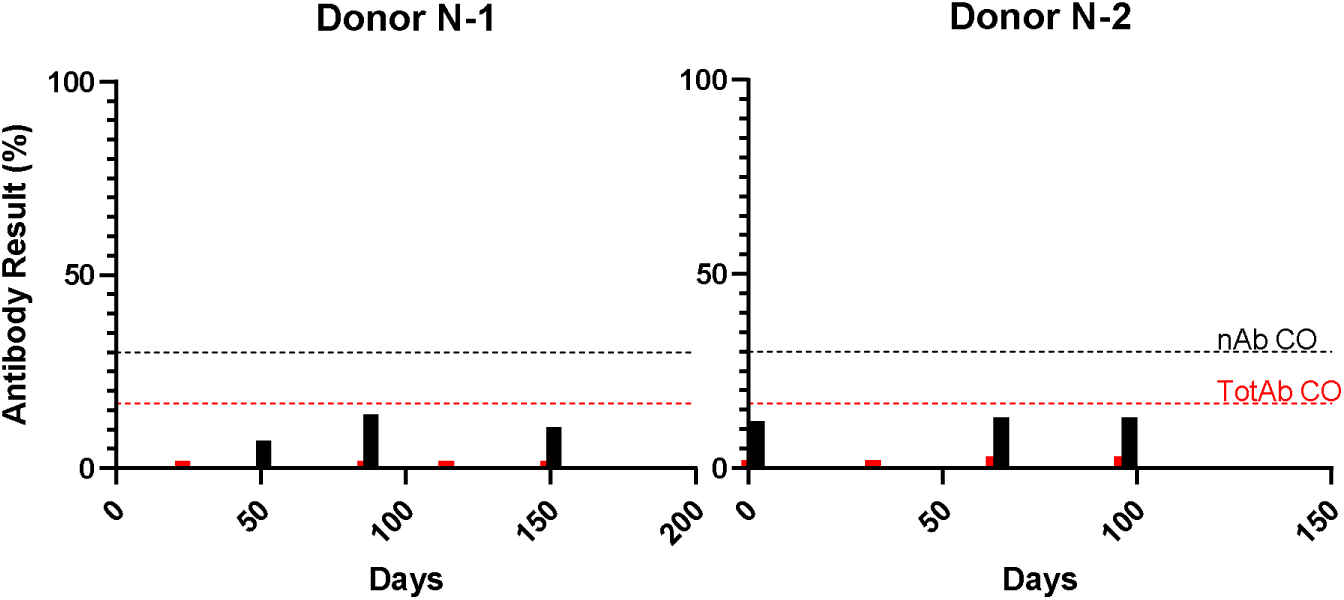
Measurements of SARS-CoV-2 total antibodies and neutralizing antibodies in two negative control donors never vaccinated nor infected with SARS-CoV-2. Neutralizing antibody levels (black bars) are reported as % signal inhibition (quenching assay) with levels ≥30% indicating a positive result [9,10] as shown by the black horizontal dashed line (nAb CO), whereas the specimen-to-cutoff ratio for the total antibody test (red bars) has been converted to % of maximum measurable specimen-to-cutoff ratio of 6.0. The cutoff for the total Ab test is shown by the red horizontal dashed line (TotAb CO). Abbreviations: Ab, antibody; CO, positive cutoff; nAb, neutralizing antibody; TotAb, total antibody.

To further understand the quantitative levels of Abs being measured by the two antibody assays, dilutions of the Human SARS-CoV-2 Serology Standard, which was calibrated to the WHO International Standard, were measured (**Figure 4**). The cPass nAb assay cutoff of ≥30% signal inhibition corresponds to approximately 1:200 dilution of the Standard, or approximately 41 IU/mL (based on the assigned potency for functional activity (neutralizing unitage) as calibrated to the WHO SARS-CoV-2 Serology International Standard). The relationship of the cPass nAb signal inhibition results to the potency of the Standard is shown in Table 1. Similarly, the Platelia total Ab assay cutoff of a specimen-to-cutoff ratio of ≥1.0 corresponds to approximately 1:200 dilution of the Standard.

**Figure 4.**
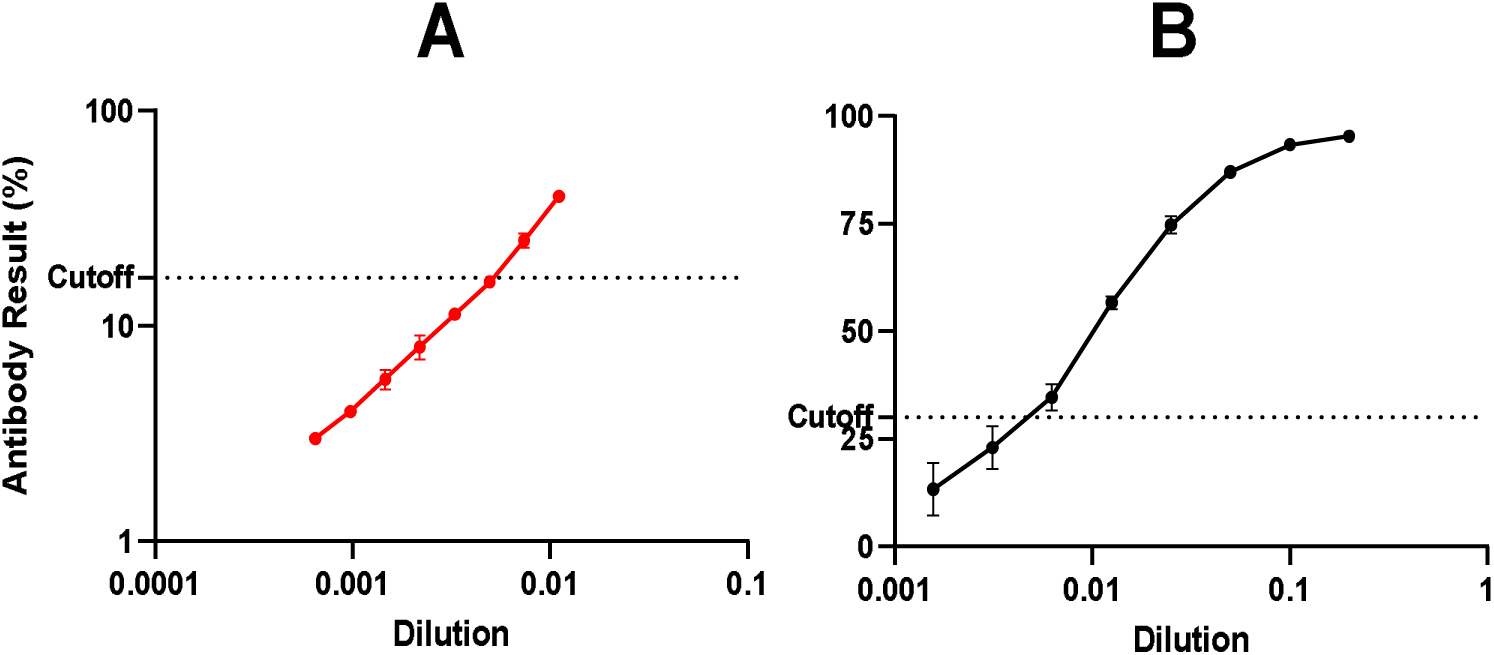
Measurements of SARS-CoV-2 total antibodies (A) and neutralizing antibodies (B) in the Human SARS-CoV-2 Serology Standard run in an 8-point dilution series. For the total Ab assay, the Standard was diluted at 1:90, followed by a 1.5-fold dilution series; for the nAb assay, the Standard was diluted at 1:5, followed by a 2-fold dilution series. Neutralizing antibody levels are reported as % signal inhibition (quenching assay), whereas the specimen-to-cutoff ratio for the total antibody test has been converted to % of maximum measurable specimen-to-cutoff ratio of 6.0. The respective cutoff for each assay is shown as a horizontal dashed line.

**Table 1.** Relationship of cPass nAb signal inhibition results to the assigned potency for functional activity of the Human SARS-CoV-2 Serology Standard (calibrated to the WHO International Standard).

| **% Signal Inhibition** | **Potency (IU/mL)** |
| --- | --- |
| 13% | 13 |
| 23% | 25 |
| 35% | 51 |
| 57% | 102 |
| 75% | 203 |
| 87% | 407 |
| 93% | 813 |
| 95% | 1626 |

Note: Percent signal inhibition values $\geq 30\%$ are reported positive based on previously conducted clinical validation studies<sup>7</sup>.

### 3.2 SARS-CoV-2 RBD specific T cell Responses

We selected 11 donors for serial testing with the T cell restimulation assay based on their infection and immunization status. In all cases, the controls performed as expected; IFN-γ concentrations for the Nil (negative) controls were ≤0.02 IU/mL, and concentrations for the Mitogen (positive) controls were greater than those measured for the corresponding donor’s SARS-CoV-2-stimulated specimens and ranged from 2.7-18.6 IU/mL. Generally, donors who were previously infected showed heterogeneous but typically strong cell-mediated responses, while those vaccinated and not previously infected showed a measurable but on average lower T cell response (**Figure 5**). Specifically, three donors who were previously infected and subsequently vaccinated, including donor I-1 who is taking an immunosuppressant, showed very strong T cell-mediated responses, while the other 4 donors who were previously infected and subsequently vaccinated showed more modest T cell responses (**Figure 5**). In the latter cases, the responses were comparable to that observed for the three vaccinated and not previously infected donors. The specimen from the donor who was neither immunized nor infected showed reactivity comparable to the Nil control indicating high specificity of the assay.

**Figure 5.**
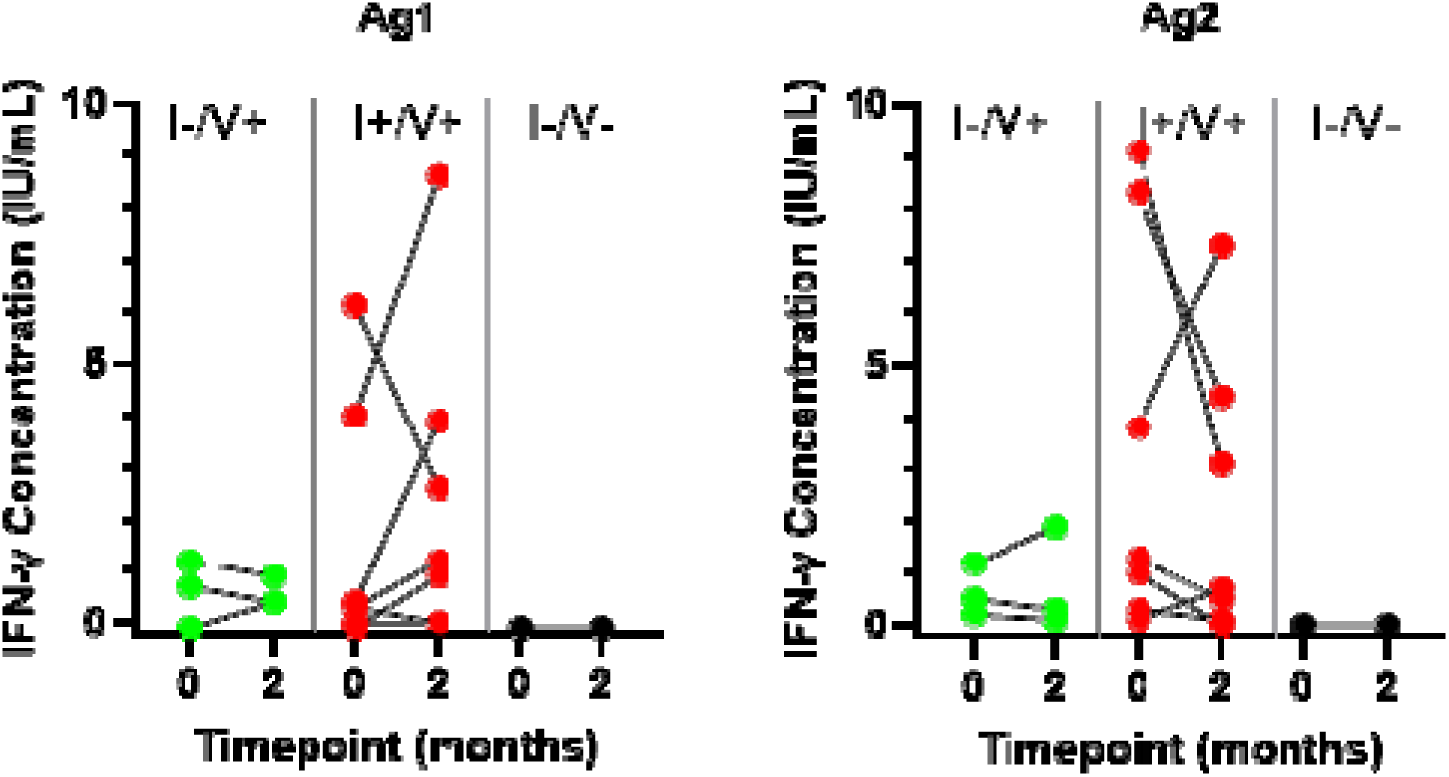
Measurements of IFN-γ release by T cells to SARS-CoV-2 peptides (Ag1 & Ag2) 2 months apart in a subset of case study donors (n=11): 3 donors were vaccinated and not previously infected (I-/V+, green), 7 were infected and subsequently vaccinated (I+/V+, red), and 1 donor was neither vaccinated nor previously infected (I-/V-, black). IFN-γ concentrations are reported in international units (IU/mL).

## 4. Discussion

Together, the data presented illustrate the sensitivity and specificity of several novel assays in monitoring critical immune responses to SARS-CoV-2 infection and/or vaccination, particularly at or near the positive to negative threshold, where the clinical utility of these tests as biomarkers may be most meaningful. Significantly, specific T and B cell activity elicited from either previous infection and/or vaccination could be monitored serially in a standard clinical laboratory setting and with standard testing technologies. Critical features of these *in-vitro* assays also include their non-invasiveness and low-cost shipping requirements. In the case of the neutralizing antibody assay, it has demonstrated excellent clinical concordance with both live virus and pseudovirus assays and is more cost-effective, offers rapid turn-around^5^ and is scalable with automation^13^. The neutralization antibody assay is unique as well, since it is the only functional assay of antibodies to the SPIKE RBD that is FDA emergency use authorized. Such technology could enable large-scale, highly specific monitoring of functional anti-SARS-CoV-2 antibodies, should such surveillance become advisable, particularly for individuals at high-risk of severe disease, such as the immune-compromised.

Results obtained for the NCI Human SARS-CoV-2 Serology Standard tested on both the Platelia Total Ab assay and the cPass Neutralization Ab assay suggest results for both assays may be quantified relative to the WHO International Standard. Global harmonization studies are underway to elucidate correlations between this Standard and others as available using these assays.

Since antibody titers alone do not present a complete picture of adaptive immunity, a novel blood cell-mediated T cell response assay was included in this observational study. Like the neutralizing and total antibody assays, the SARS-CoV-2 RBD T cell assay utilizes a readily available ELISA kit, can be automated and is cost-effective, since it requires no cell culture. Similar to the trends observed with nAb, some individuals with past natural infection and subsequent vaccination demonstrated high T cell activity while others had more modest T cell activity comparable to those individuals who had been vaccinated and never infected. However, responses were heterogeneous. Donor I-1 who was taking an immunosuppressant maintained high levels of nAb and T cell activity for 5 months post-vaccination, while other immunosuppressed donors have not shown such levels of nAb.

We note that our findings of more significant and sustained spike-specific T cell responses in previously infected/vaccinated individuals as compared to infection-naïve/vaccinated individuals are consistent with findings from other studies^14^. Immune responses to specific variants of SARS-CoV-2 and to individuals who have received booster doses of vaccines are also under investigation. Finally, while some studies to date have attempted to correlate serological antibody titer decline with breakthrough infection^15^, research to determine what levels of antibody and/or T cell responses may confer protective immunity is still ongoing^14,16^.

In summary, the case study data in this observational study represent snapshots for individuals who have been SARS-CoV-2 infected and/or vaccinated. This study adds to the population-based clinical studies, and the heterogeneous neutralizing antibody and T cell responses illustrate the value of testing on an individual basis in both the immunocompetent and immunocompromised. The development of robust and reliable biomarkers and associated clinical tests that can be used as correlates of protective immunity after SARS-CoV-2 infection and/or immunization may help to inform appropriate protective measures, particularly for immunocompromised and vaccine insensitive individuals. Further, studies demonstrating vaccine potency have increasingly used neutralizing antibody assays as surrogate measures of potency^17^. We have presented, in various cases studies, examples of how rapid and scalable in vitro tests that measure B and T cell functional immunity to SARS-CoV-2 may offer insights into the duration and degree of potentially protective immunity. Additionally, we have provided evidence to suggest that both B cell assays, nucleocapsid protein and neutralizing, may be quantified by calibrating results to a well-characterized International Standard^18^. This relationship has the potential to allow comparisons of the cPASS nAb test measurement in future studies intended to evaluate correlates of protection. Although assaying for protective immunity after infection or vaccination is not recommended for the general population at this time, greater immune surveillance incorporating these technologies could broadly inform vaccination and booster recommendations especially to the populations at risk for SARS-CoV-2 infections, severe disease, and hospitalization.

## Data Availability

All data produced in the present work are contained in the manuscript

## Acknowledgments

The authors recognize and are grateful for the support of our study donors. Coordination of test logistics and phlebotomy services were provided by Raynie Baca, Peter Plankis, Micah Bruns and their teams at Biodesix, Inc. Technical collaborative discussions were held with teams at Bio-Rad, Inc. and GenScript, Inc. Medical writing services were provided by Rachel Hartfield, PhD and Ubaradka G. Sathyanarayana, PhD and supported by Biodesix, Inc.

## Author Contributions

Conceptualization, L.P., A.P., N.G., T.J.W and G.P.; methodology, L.P., A.P., B.D., L.J., A.W., N.G., and G.P.; formal analysis, L.P. and G.P.; investigation, L.P., A.P., B.D., N.G., and G.P.; data curation, L.P.; writing original draft preparation, L.P and G.P.; writing review and editing, L.P., A.P., N.G., B.D., A.W., L.J., T.J.W and G.P.; supervision, L.P. and G.P; administration, N.G and G.P. All authors have read and agreed to the submitted version of the manuscript.

## Institutional Review Board Statement

The observational study was conducted according to the guidelines of the Declaration of Helsinki and approved by the Advarra Institutional Review Board (BDSX-CD-004, 12 March 2021).

## Data Sharing Statement

The data that supports the findings of this study are available on request.

